# Safety, Tolerability, and Immunogenicity of COVID-19 Vaccines: A Systematic Review and Meta-Analysis

**DOI:** 10.1101/2020.11.03.20224998

**Authors:** Ping Yuan, Pu Ai, Yihan Liu, Zisheng Ai, Yi Wang, Weijun Cao, Xiaohuan Xia, Jialin C. Zheng

## Abstract

We aimed to summarize reliable medical evidence by the meta-analysis of all published clinical trials that investigated the safety, tolerability, and immunogenicity of vaccine candidates against coronavirus disease 2019 (COVID-19), caused by severe acute respiratory syndrome coronavirus 2 (SARS-CoV-2). The PubMed, Cochrane Library, EMBASE, and medRxiv databases were used to select the studies. 7094 articles were identified initially and 43 were retrieved for more detailed evaluation. 5 randomized, double-blind, placebo-controlled trials were selected. A total of 1604 subjects with either vaccines or placebo infections were included in the meta-analysis within the scope of these articles. According to the results, there is an increase in total adverse events for subjects with either low (95% *CI*: 1.90-4.29) or high (*CI*: 2.65-5.63) dose vaccination. The adverse effects of COVID-19 vaccine are mainly local ones including pain, itching, and redness, and no significant difference was identified in the systemic reactions. All adverse effects were transient and resolved within a few days. Moreover, the neutralizing and IgG antibody levels post different dose vaccinations were all significantly increased at day 14/21 (*P* = 0.0004 and *P* = 0.0003, respectively) and day 28/35 (*P* < 0.00001) in vaccine groups compared to placebo controls. Besides, the levels of neutralizing and IgG antibodies were also elevated significantly at from day 14 to 35, versus day 0 (All *P* < 0.001). In conclusion, our analysis suggests that the current COVID-19 vaccine candidates are safe, tolerated, and immunogenic, which provides important information for further development, evaluation, and clinical application of COVID-19 vaccine.

## Introduction

Coronavirus disease 2019 (COVID-19), caused by severe acute respiratory syndrome coronavirus 2 (SARS-CoV-2), has rapidly spread globally [1]. On 11 March 2020, the World Health Organization (WHO) declared the COVID-19 outbreak a pandemic. As of November 3^rd^, 2020, there have been more than 46 million confirmed cases of COVID-19 and more than 1 million deaths worldwide. SARS-COV-2 infected individuals experience a wide range of symptoms include fever, cough, shortness of breath, and fatigue [2]. COVID-19 patients with severe pneumonia have acute respiratory distress syndrome (ARDS) and changes in heart and liver function as a secondary or related consequence of disease, which could lead to multiple organ failures and death [3].

Vaccine is one of the best armamentaria in public health especially where no effective treatment is available against infectious disease. Due to the global pandemic, the vaccines are urgently needs to protect people against COVID-19. To date, more than 120 COVID-19 preclinical candidate vaccines have been developed globally, and 10 of them are in human trials (https://www.who.int/emergencies/diseases/novel-coronavirus-2019/covid-19-vaccines). The safety, tolerability, and immunogenicity results of the phase 1/2 trials of multiple vaccines, including RNA-based vaccines (e.g. BNT162b1/2 and mRNA-1273) and adenovirus-vectored vaccines (e.g. ChAdOx1 nCoV-19 and non-replicating adenovirus type-5/26), have been published or made available on preprint servers [4-6]. In addition, there are a great amount of vaccine candidates, such as a series of DNA vaccines and inactivated vaccine BBIBP-CorV, that have been reported to protect non-human primates against SARS-CoV-2 with varying efficacy [7, 8].

To our knowledge, no comprehensive meta-analysis has been published focusing on the safety, tolerability, and immunogenicity of current COVID-19 vaccine candidates, although numbers of basic research and clinical trial results have been announced. Our study will be the first one summarizing and analyzing the aforementioned clinical findings of current vaccine candidates to acquire more accurate conclusions, providing important guidance for the development and clinical application of COVID-19 vaccine.

## Methods

### Search strategy

The study was designed according to the standards set forth by the PRISMA (Preferred Reporting Items for Systematic Reviews and Meta-Analyses) statement [9, 10]. We searched systematically in PubMed, Embase, the Cochrane Library, and medRxiv using the key words “COVID-19” or “SARS-CoV-2” and “vaccination” or “vaccine” to identify all published and pre-publication studies from the database inception to October 20, 2020. The search is limited in English language papers.

### Study selection

We included randomized, double-blind, placebo-controlled studies. Study inclusion criteria were: healthy adults (18 years and older) without history of SARS-CoV (via on-site inquiry) or SARS-CoV-2 infection (via serological and nucleic acid test); Before vaccination or placebo and at days 14, 21, 28 or 35 after injections, subjects were asked to record any injection local adverse reactions (e.g. pain, itching, redness, swelling and induration) and systemic adverse reactions (e.g. coughing, diarrhea, fatigue, fever and headache), and subjects received routine laboratory blood tests and antibody tests included the neutralizing antibody and the specific IgG-binding antibody titers.

Injections of the viral vaccines [1× 10^11^/ mL or 5× 10^1^□/ mL Ad5 vectors-viral particles, or 2.5, 5, and 10 μg antigen protein content per dose SARS-CoV-2 strain vaccine, or 5 μg and 25 μg doses of rSARS-CoV-2 with or without Matrix-M1 adjuvant (50 μg dose), or 3 μg/0.5 mL and 6 μg /0.5mL inactivated SARS-CoV-2 whole virion], RNA vaccine (10 μg, 30 μg and 100 μg per dose), or placebo, were given to participants intramuscularly in the arm.

Reviews, editorials, letters, animal studies, case reports, commentaries, reviews and conference abstracts were excluded. Studies were excluded if there was an overlap in subjects with another study within the same analysis. Thus, if some subjects could possibly have been included in different phase trial, they were only included once in any given analysis. Therefore, there was no overlap in populations included in our meta-analyses.

### Data extraction and quality assessment

Our data was extracted by three investigators (P.Y., P.A., Y.L.) independently, and disagreements were resolved by their consensus or consultation with the third author. For each study, we collected the journal, date of publication, first author, sample size, study design, population demographics (including ethnicity, sex ratio, and age range or mean age), the number of vaccinated and unvaccinated subjects, outcomes, adverse reaction, and adjusted measures of the corresponding 95 % confidence intervals (*CI*).

Risk of bias was assessed for the domains suggested by the Cochrane Handbook of Systematic Reviews [11], specifically emphasizing sequence generation, allocation concealment, blinding, outcomes assessment, and selective reporting for the 5 included trials. We did not detect clear publication bias, because the number of included studies was small.

### Outcomes

Main outcomes measures of our meta-analysis included safety and immunogenicity of the vaccine. According to the adverse reaction extracted from the original studies, the safety outcomes were evaluated as individual conditions including local adverse reactions (pain, itching, redness, swelling, induration) and systemic adverse reactions (coughing, diarrhea, fatigue, fever, headache, nausea and vomiting, pruritus, muscle pain, joint pain and malaise or anorexia). Similarly, immunogenicity outcomes were evaluated by abnormal hemoglobin, alanine aminotransferase and total bilirubin, IgG or other specific antibody responses to the receptor binding domain, and neutralizing antibodies to live SARS-CoV-2.

### Data synthesis and statistical analysis

For 5 included studies, the differences in frequency of adverse events and change of antibody levels and laboratory parameters with vaccination versus placebo or baseline were pooled, stratified across studies, and analyzed using random-effects or fixed effect models with inverse variance weighting. Random-effects models were used when the I^2^ statistics to estimate the proportion of variation attributable to between-study heterogeneity more than 50% or p < 0.1. And fixed effect models were used when I^2^ less than 50% or *P* > 0.5. Separate models were constructed for low and high dose vaccination, 14/21 and 28/35 days of follow-up. The magnitude of heterogeneity was estimated using the I^2^ statistic, an estimate of the proportion of the total observed variance that is attributed to between-study variance.

Pooled effects on adverse events and abnormal hemoglobin, alanine aminotransferase and total bilirubin levels were presented as odds ratio (OR) with corresponding 95% *CI*. Pooled effects on antibody levels were presented as weighted mean differences with corresponding 95% *CI*. We considered *P* < 0.05 significant. Throughout, values were presented as mean ± SD unless otherwise stated. Additionally, we used a funnel plot to evaluate the publication bias. Analyses were performed using the Cochrane Collaboration Review Manager (version 5.2, Cochrane Collaboration, Copenhagen, Denmark).

## Results

### Characteristics of the Studies

Of 4223 articles from PubMed, 187 articles from the Cochrane Library, 2684 articles from EMBASE, and 2736 from medRxiv identified initially, 43 were retrieved for more detailed evaluation, Subsequently, 5 studies that included 1604 subjects were finally included in the analyses (**Figure 1**). The baseline characteristics of the included 5 randomized, double-blind, placebo-controlled trials including the design and methods were summarized (**Table 1**). Studies included subjects with healthy adults over 18 years old. The vaccination procedures were similar in these studies, including the injection site, the period of follow-up, primary and secondary end-point events, and the recorded adverse reactions. Three studies designed a single injection and two studies designed two to three vaccinations. The included 5 studies utilized a range of vaccine doses from 2.5 μg to 100 μg per dose with at least two to three dose gradients. The risk of bias was low in the majority of studies, with a detailed assessment available (**Figure 2**).

**Table 1.** Baseline characteristics of all included studies.

| Table 1 Baseline Characteristics of All Included Studies |  |  |  |  |  |  |  |
| --- | --- | --- | --- | --- | --- | --- | --- |
| Study<br>First Author<br>(Ref.) | Type of Study | Placebo/<br>Vaccine<br>Subjects<br>n | Subjects<br>Characteristi<br>cs | Adverse event |  | Vaccines Characteristics | Duration and Follow up |
|  |  |  |  | Local<br>reactions | Systemic<br>reactions |  |  |
| Randomized Control Trial |  |  |  |  |  |  |  |
| Xia 2020 | randomized<br>double-blind<br>placebo-controlled<br>studies | 80/240 | asain<br>mean age,<br>42.8 years<br>male, 37.5% | pain<br>itching<br>redness<br>swelling | fever, coughing<br>diarrhea, fatigue<br>headache, pruritus<br>nausea, vomiting | investigational inactivated whole-virus COVID-19 vaccine;<br>phase 1 trial: 2.5, 5, and 10 µg/dose<br>intramuscular injections;<br>phase 2 trial: 5 µg/dose in 2 schedule groups<br>intramuscular injections;<br>WIV04 strain, National Genomic Data Center of the Chinese Academy of Science accession No. SAMC133237, and GenBank accession numberMN9 | phase 1 trial:<br>4,14, 21 days after each injection in first dose;<br>4 days and 14 days in second and third dose;<br>phase 2 trial:<br>14 days after second dose |
| Zhu 2020 | randomized<br>double-blind<br>placebo-controlled<br>studies | 126/382 | asain<br>mean age,<br>39.7 years;<br>male, 50% | pain<br>Induration<br>redness<br>swelling<br>itch | fever, headache, fatigue,<br>vomiting diarrhoea, cough, hypersensitivity, nausea,<br>muscle pain, joint pain, dyspnoea, appetite impaired,<br>oropharyngeal pain, pruritus,<br>syncope, mucosal abnormality | Ad5-vectored COVID-19;<br>A single injection of the vaccines of 1×10<sup>11</sup> or 5×10<sup>11</sup> viral particles per mL;<br>Wuhan-Hu-1, GenBank accession number YP_009724390 | 14 and 28 days after each injection |
| Keech 2020 | randomized<br>double-blind<br>placebo-controlled studies | 23/108 | mean age,<br>30.8 years;<br>male, 50.4% | pain<br>erythema redness<br>induration swelling<br>tenderness | arthralgia, fatigue,<br>fever, headache,<br>myalgia, nausea,<br>malaise | NVX-CoV2373<br>one injection on day 0 and one on day 21<br>(GenBank accession number, MN908947; nucleotides 21563–25384) | 7, 21, 28, and 35 days after each injection |
| Zhang 2020 | randomized<br>double-blind<br>placebo-controlled studies | 120/480 | mean age,<br>42.09 years ;<br>male, 47.2% | pain<br>induration<br>swelling<br>redness<br>rash<br>pruritus | fever, diarrhea, cough, fatigue<br>acute allergic reaction, vomiting,<br>abnormal skin and mucosa,<br>anorexia, nausea, headache,<br>muscle pain, | SARS-CoV-2 inactivated vaccine (CoronaVac);<br>3 µg/0.5 mL or 6 µg /0.5mL | 14, 28, and 42 days after each injection |
| Mulligan 2020 | randomized<br>double-blind<br>placebo-controlled studies | 9/36 | mean age,<br>35.4years ;<br>male, 51.1% | pain<br>redness<br>swelling | fever, fatigue, headache, chills,<br>vomiting, diarrhoea, medication,<br>muscle pain, joint pain, | BNT162b1;<br>10 µg and 30 µg per dose;<br>genetic sequence MN908947.3 | 7, 14, 21, 28, and 35 days after each injection |

**Figure 1.**
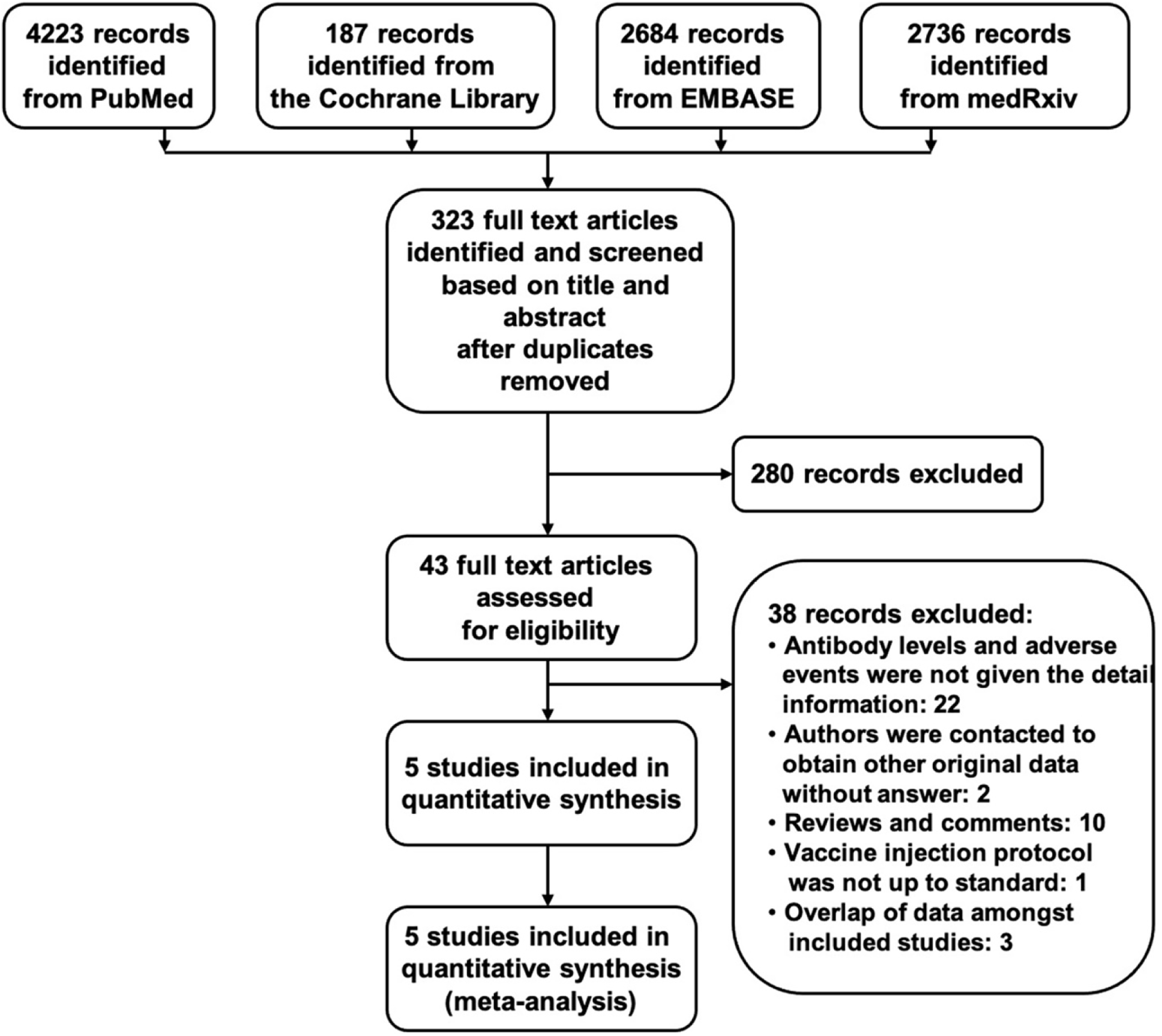
Flowchart showing the progress through the stages of meta-analysis.

**Figure 2.**
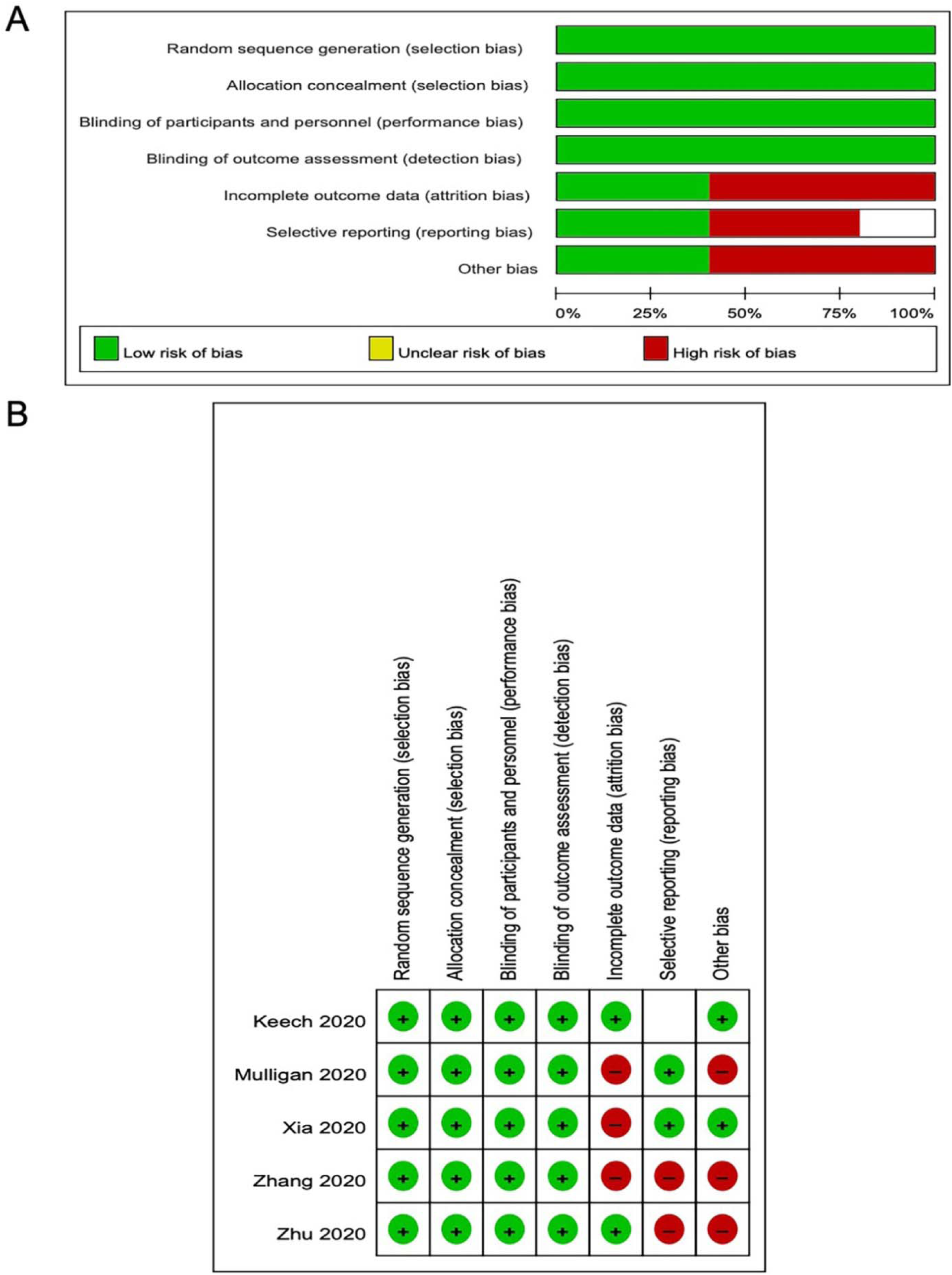
Overall risk of bias assessment using the Cochrane tool and risk of bias assessment by individual trials. A: overall risk of bias assessment using the Cochrane tool. B: Risk of bias assessment by individual trials.

### Effect of vaccination on adverse reactions

For the meta-analysis, we separated the adverse events based on vaccine vs. placebo injection as reported by individual studies. In general, we observed there was an increase in total adverse events for subjects with low dose vaccine injection [OR: 2.86; 95% *CI*: 1.90-4.29, *P* < 0.00001] (**Figure 3**). Especially, the local reactions were significantly enhanced in subjects with low dose vaccine groups [OR: 2.07; 95% *CI*: 1.07-4.00, *P* = 0.03] (**Figure 3**). However, the systemic reactions were no significantly changed between vaccine and placebo groups [OR: 1.28; 95% *CI*: 0.67-2.43, *P* = 0.46]. There were high amounts of heterogeneity about total adverse events and local reactions and a lower amount of heterogeneity of systemic reactions in these studies (I^2^ = 79%, 73%, and 10%; **Figure 3**).

**Figure 3.**
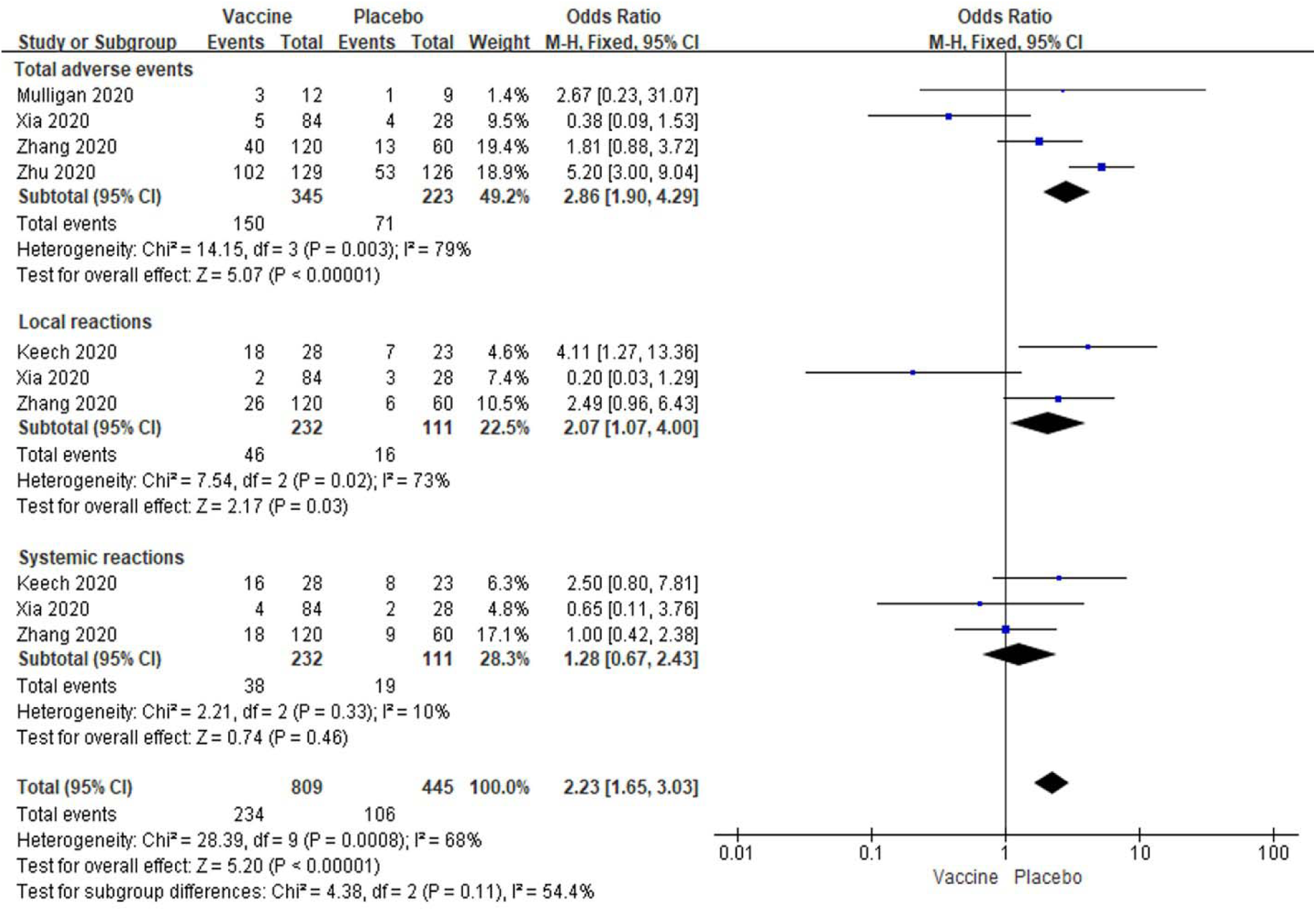
Meta-analysis of effect of low dose vaccine on adverse reactions between vaccine and placebo groups.

The similar data indicated total adverse events [OR: 3.86; 95% *CI*: 2.65-5.63, *P* < 0.00001] and local reactions [OR: 4.80; 95% *CI*: 2.36-9.78, *P* < 0.00001] were significantly increased in subjects with high dose vaccine injection compared to placebo injection (**Figure 4**). And the systemic reactions also show no significant alteration in subjects with high dose vaccination [OR: 1.81; 95% *CI*: 0.91-3.59, *P* = 0.09] (**Figure 4**). Modest to high amounts of heterogeneity were showed in analyses of total adverse events, local reactions and systemic reactions (I^2^ = 53%, 53%, and 80%; **Figure 4**).

**Figure 4.**
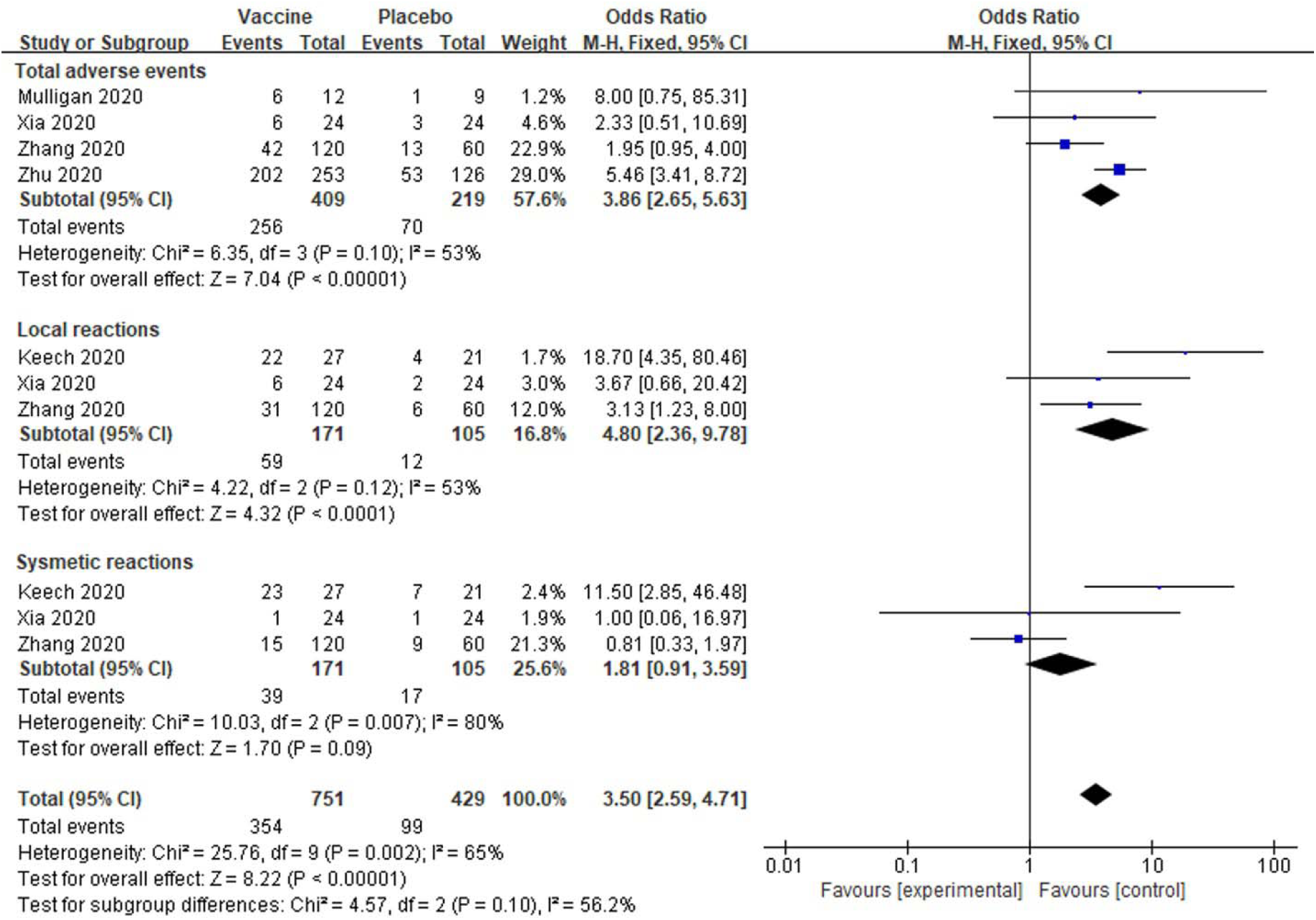
Meta-analysis of effect of high dose vaccine on adverse reactions between vaccine and placebo groups.

In low dose vaccine subgroup analyses showed total local events were significantly higher in vaccine groups than placebo groups [OR: 2.72; 95% *CI*: 1.36-5.47, *P* = 0.005] (**Figure 5**). However, any local reactions, including pain, itching, redness, swelling and induration, were no significantly changed between the two groups (All *P* > 0.05, **Figure 5**). There were low to modest amounts of heterogeneity about all local reactions besides pain among these studies (I^2^= 86%, 0%, 12%, 0% and 48%; **Figure 5**). In analysis of systemic reactions, we have only analyzed some common systemic reactions in all studies. There were significantly increased of fatigue, headache, muscle pain, joint pain and malaise/anorexia in subjects with low dose vaccine injections compared to placebo injections (All *P* < 0.05, **Figure 6**). There were no significantly changed in coughing, diarrhea, fever, pruritus, nausea and vomiting in vaccine groups with lose dose compared to placebo groups (All *P* > 0.05, **Figure 6**). There were also low to modest amounts of heterogeneity about all systemic reactions (I^2^ = 0% to 51%; **Figure 6**).

**Figure 5.**
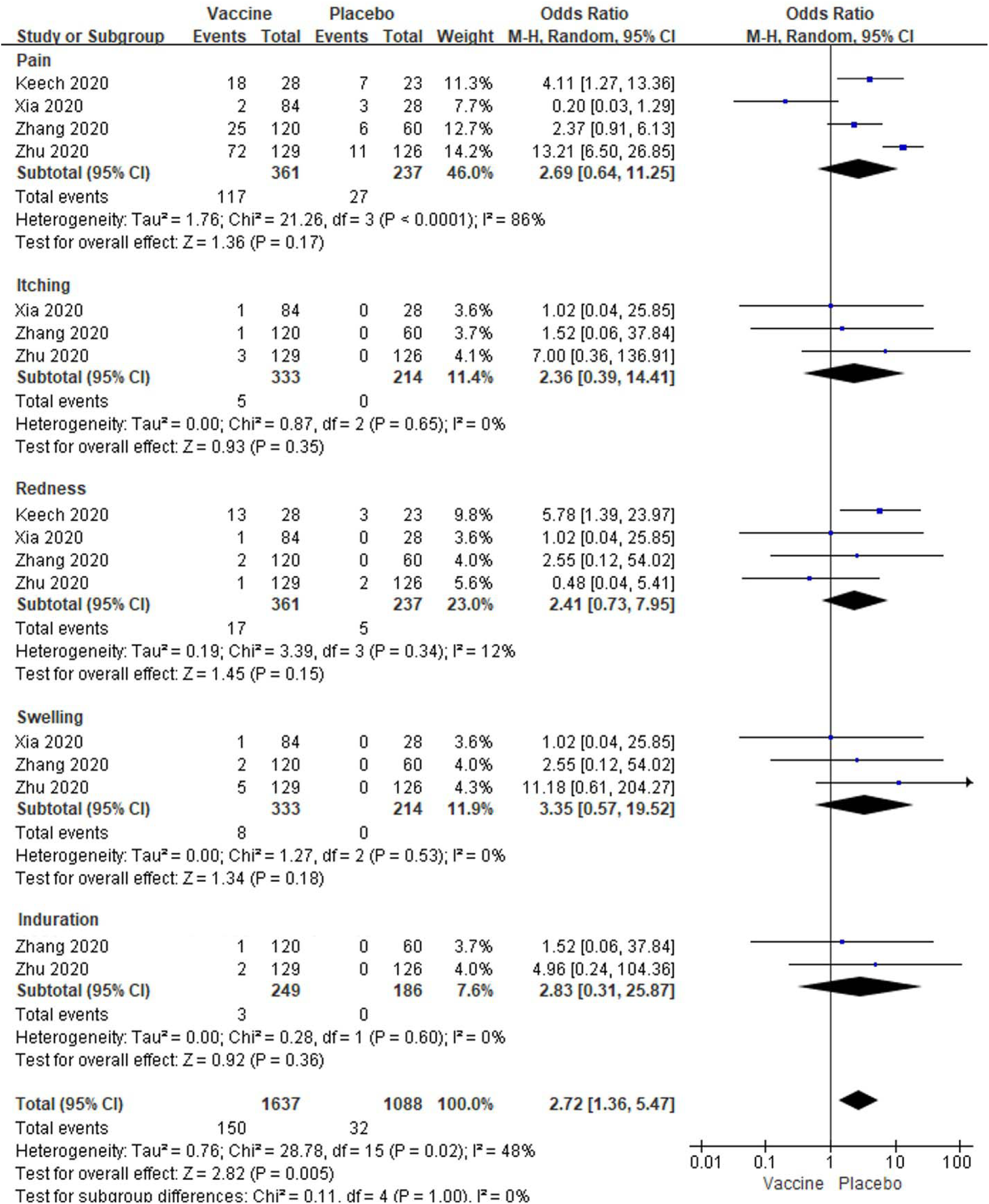
Meta-analysis of effect of low dose vaccine on local adverse reactions between vaccine and placebo groups.

**Figure 6.**
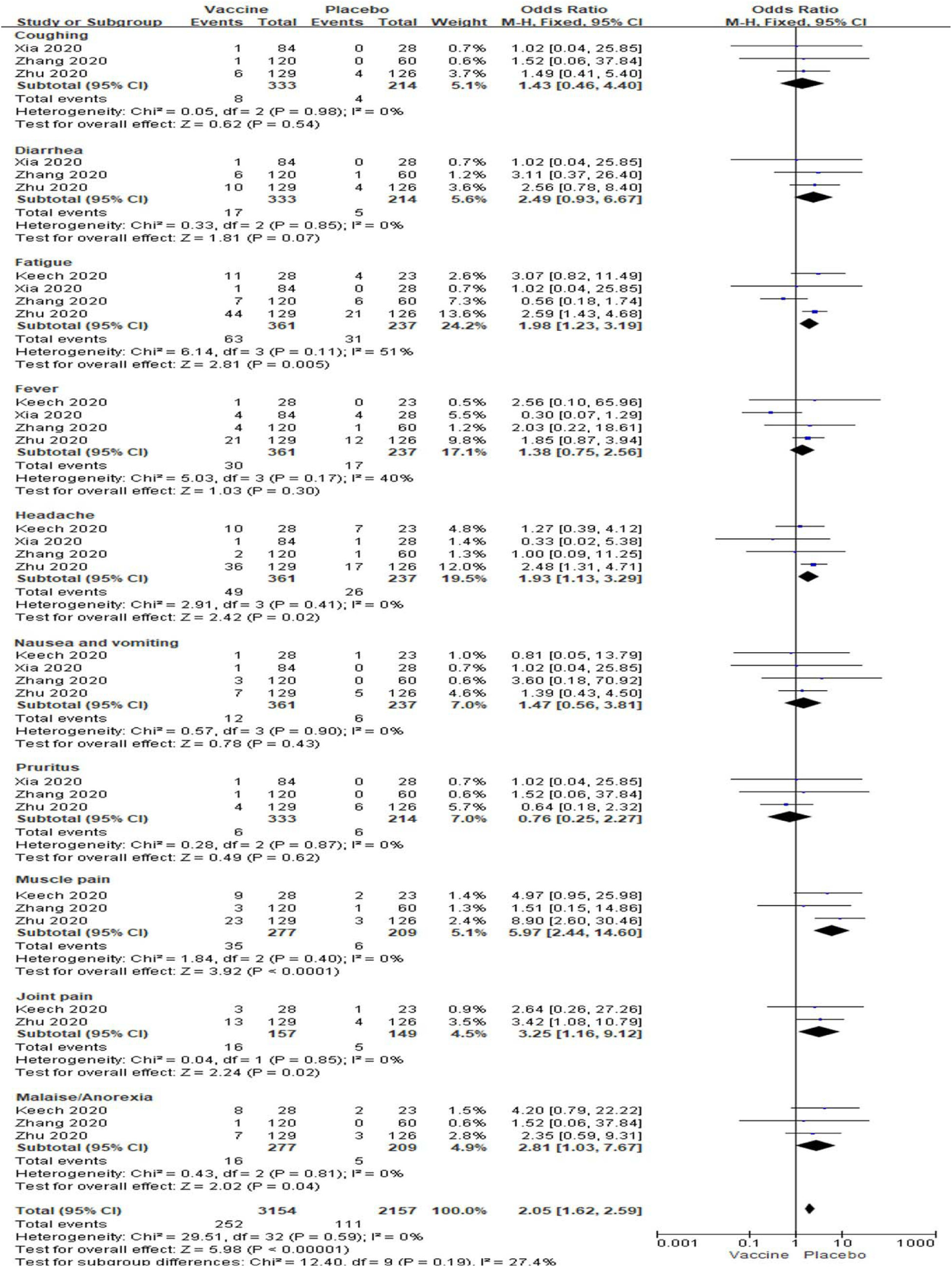
Meta-analysis of effect of low dose vaccine on systemic adverse reactions between vaccine and placebo groups.

In high dose vaccine subgroup analyses, total local events, pain, itching and swelling were also significantly enhanced in vaccine groups (All *P* < 0.05, **Figure 7**), but redness and induration was no changed between the two groups [OR: 2.57; 95% *CI*: 0.76-0.72, *P* = 0.13; OR: 1.51; 95% *CI*: 0.16-14.64, *P* = 0.72] (**Figure 7**). Low to high amounts of heterogeneity were showed in the analyses of systemic reactions (I^2^ = 0% to 67%; **Figure 7**). Total systemic event, fatigue, fever, headache, nausea and vomiting, muscle pain, joint pain and malaise/anorexia (All *P* < 0.05) besides coughing, diarrhea and pruritus (All *P* > 0.05) were also significantly higher in vaccine groups than placebo groups (All *P* < 0.05, **Figure 8**). And there were no amounts of heterogeneity about all systemic reactions (All I^2^ = 0%) besides fatigue, muscle pain and total events (I^2^ = 65%, 14% and 5%;) among these studies (**Figure 8**).

**Figure 7.**
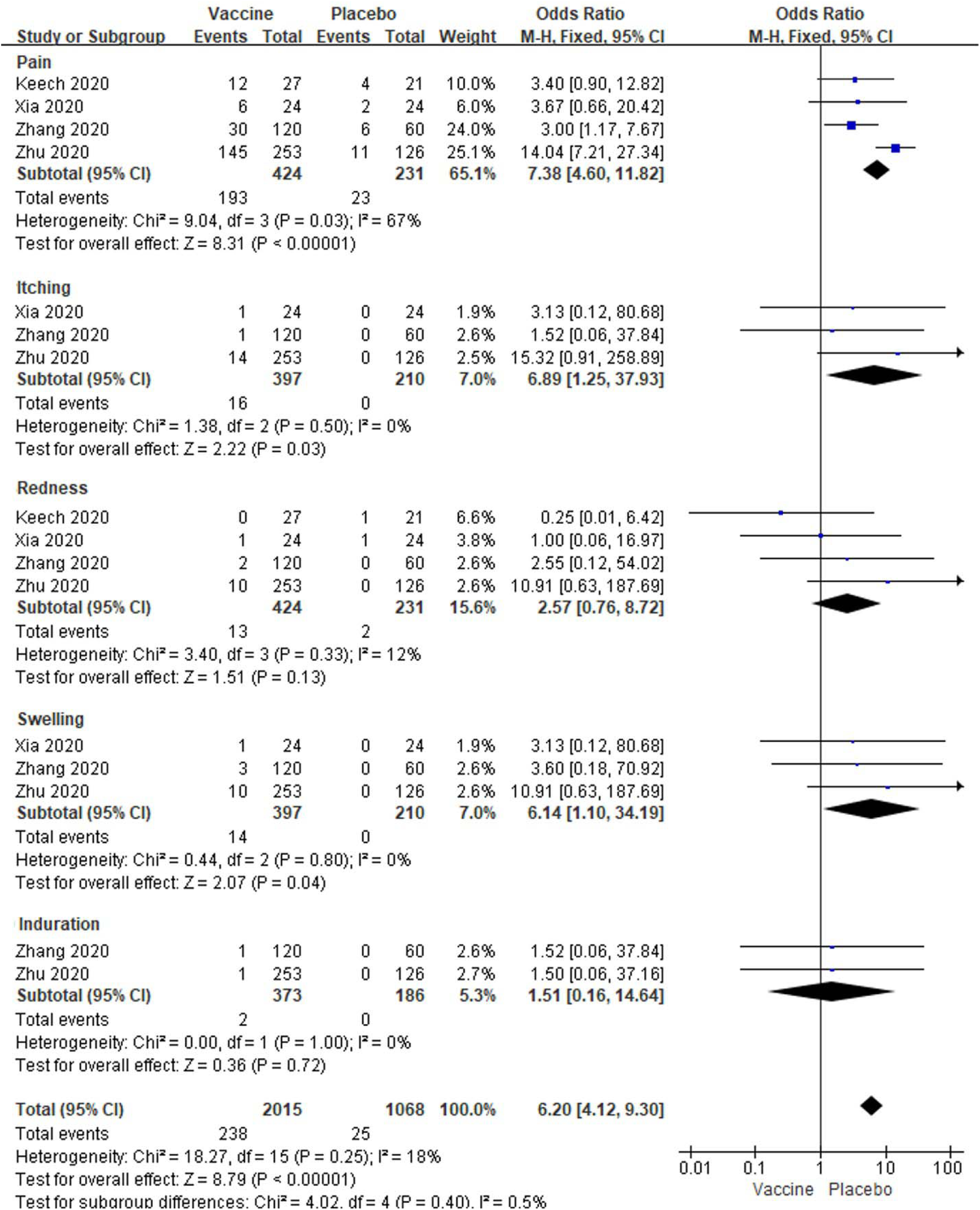
Meta-analysis of effect of high dose vaccine on local adverse reactions between vaccine and placebo groups.

**Figure 8.**
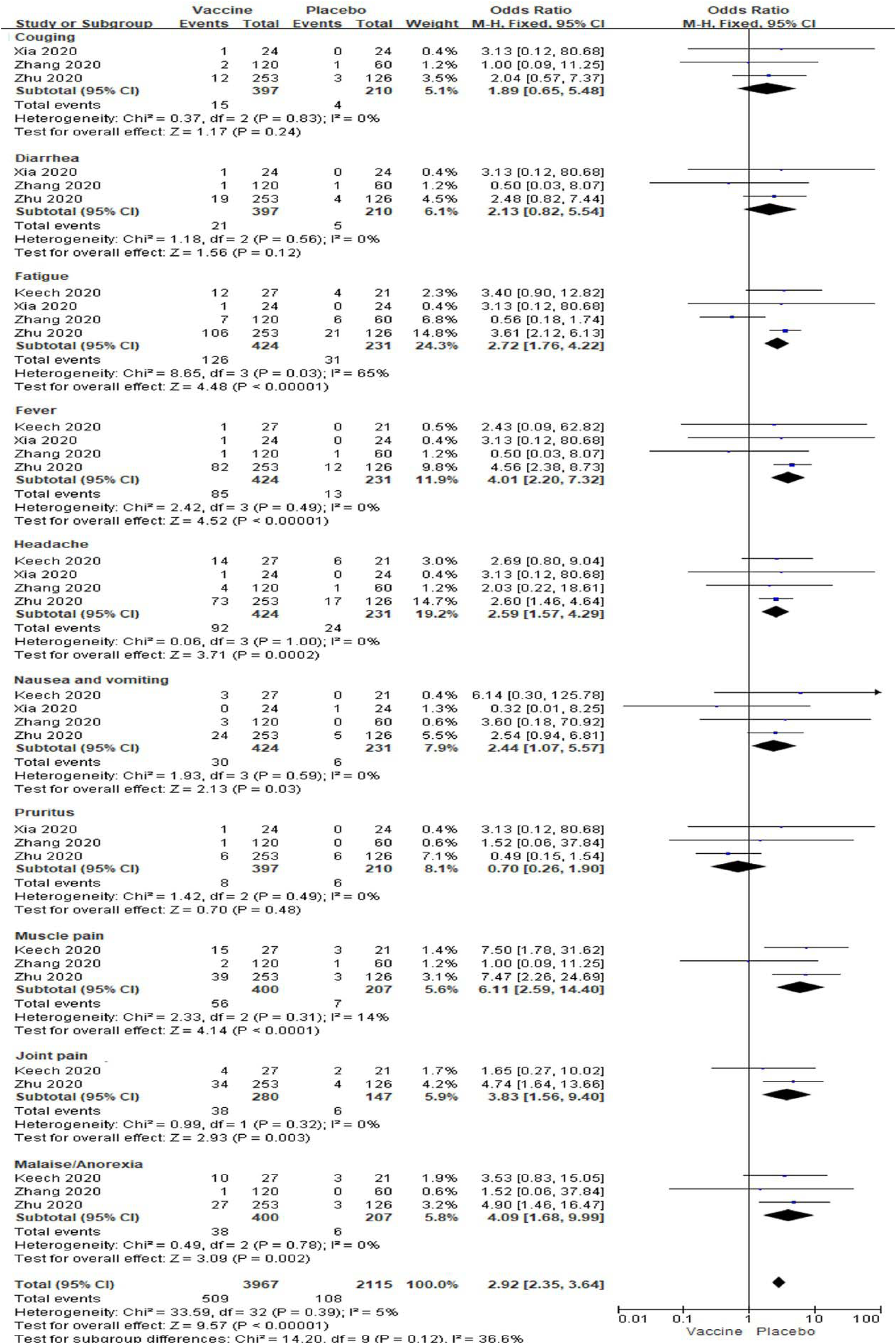
Meta-analysis of effect of high dose vaccine on systemic adverse reactions between vaccine and placebo groups.

### Effect of vaccination on neutralizing and IgG antibody responses

The neutralizing antibody levels post different dose vaccinations were all increased at day 14/21 (p = 0.0004) and day 28/35 (*P* < 0.00001) in vaccine groups compared to placebo controls, so the total neutralizing antibody levels were also significantly increased (*P* < 0.00001, **Figure 9**). Low to modest amounts of heterogeneity were showed in the analyses (I^2^ = 0%, 56% and 40 %; **Figure 9**).

**Figure 9.**
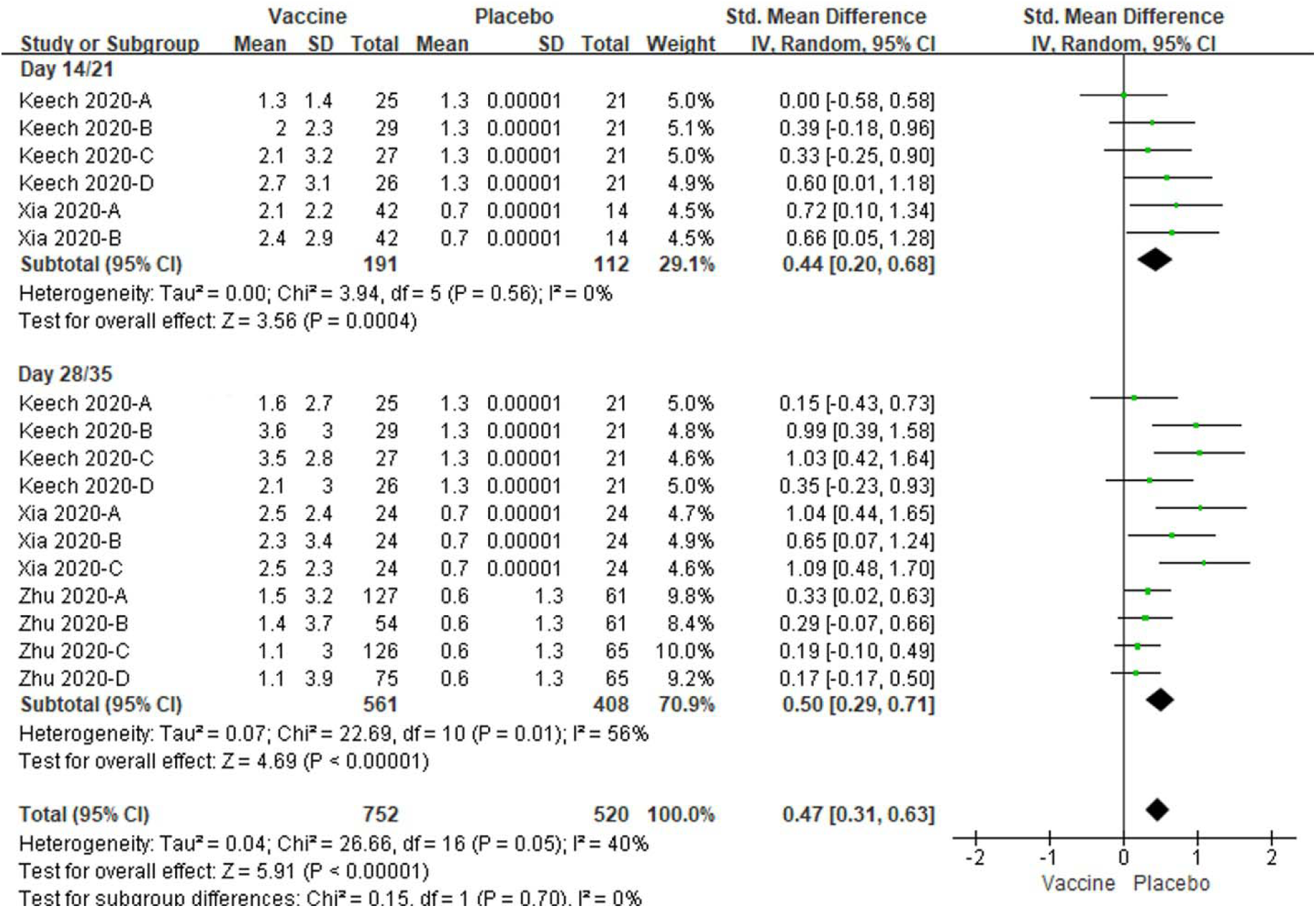
Meta-analysis of effect of vaccination on neutralizing antibody responses between vaccine and placebo groups. Keech 2020-A: 25 µg rSARS-CoV-2/0 µg Matrix-M1 on first vaccination; 25 µg rSARS-CoV-2/0 µg Matrix-M1 on second vaccination; Keech 2020-B: 5 µg rSARS-CoV-2/50 µg Matrix-M1 on first vaccination; 5 µg rSARS-CoV-2/50 µg Matrix-M1 on second vaccination; Keech 2020-C: 25 µg rSARS-CoV-2/50 µg Matrix-M1 on first vaccination; 25 µg rSARS-CoV-2/50 µg Matrix-M1 on second vaccination; Keech 2020-D: 25 µg rSARS-CoV-2/50 µg Matrix-M1 on first vaccination; 0 µg rSARS-CoV-2/0 µg Matrix-M1 on second vaccination. Xia 2020-A: low dose vaccine at day 14 or 28 follow-up, Xia 2020-B: high dose vaccine at day 14 follow-up or medium dose vaccine at day 28 follow-up, Xia 2020-C: high dose vaccine at day 28 follow-up. Zhu 2020-A: low dose vaccine and pre-existing Ad5 ≤ 200 geometric mean antibody titre. Zhu 2020-B: low dose vaccine and pre-existing Ad5 > 200 geometric mean antibody titre. Zhu 2020-C: low dose vaccine and pre-existing Ad5 ≤ 200 geometric mean antibody titre. Zhu 2020-D: low dose vaccine and pre-existing Ad5 > 200 geometric mean antibody titre.

And the specific antibody or IgG levels were significantly increased at day 14/21(*P* = 0.0003) and day 28/35 (*P* < 0.00001) in vaccine groups as well, compared to placebo controls (**Figure 10**). No amounts of heterogeneity were obtained in the analyses (All I^2^ = 0%). The total specific antibody or IgG levels were also significantly increased in vaccine groups compared to placebo controls (*P* < 0.00001, **Figure 10**). A modest amount of heterogeneity were observed in the analyses (I^2^ = 49%, **Figure 10**).

**Figure 10.**
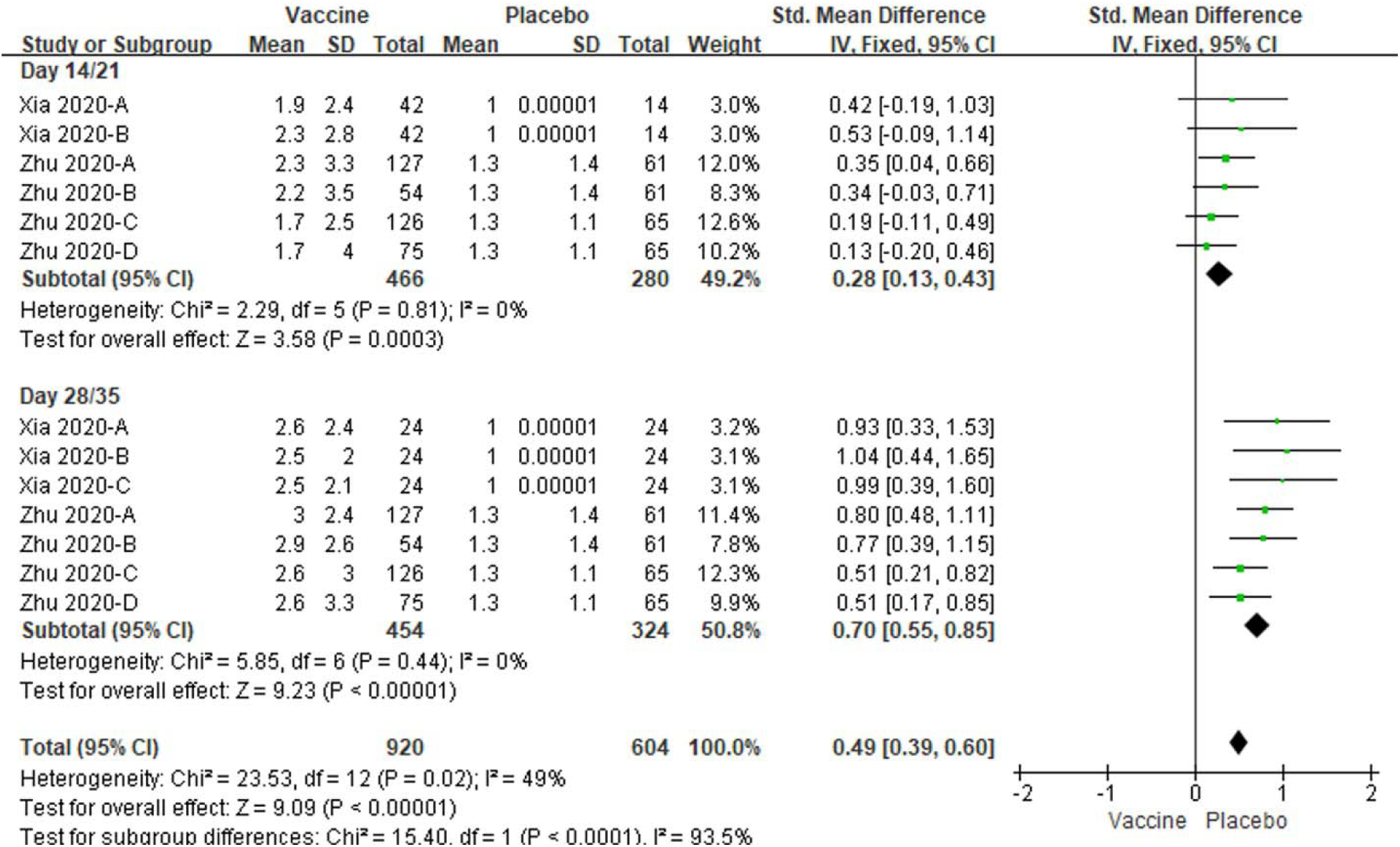
Meta-analysis of effect of vaccination on specific and IgG antibody responses between vaccine and placebo groups. Xia 2020-A: low dose vaccine at day 14 or 28 follow-up, Xia 2020-B: high dose vaccine at day 14 follow-up or medium dose vaccine at day 28 follow-up, Xia 2020-C: high dose vaccine at day 28 follow-up. Zhu 2020-A: low dose vaccine and pre-existing Ad5 ≤ 200 geometric mean antibody titre. Zhu 2020-B: low dose vaccine and pre-existing Ad5 > 200 geometric mean antibody titre. Zhu 2020-C: low dose vaccine and pre-existing Ad5 ≤ 200 geometric mean antibody titre. Zhu 2020-D: low dose vaccine and pre-existing Ad5 > 200 geometric mean antibody titre.

In the comparison between pre- and post-vaccinations, not only the levels of neutralizing antibody but also that of specific antibody or IgG were also enhanced at day 14, 21, 28 and 35, versus day 0 (All *P* < 0.001, **Figure 11, 12**). Low amounts of heterogeneity were related to both analyses, at days 14/21 and days 28/35 (I^2^ = 0% to 37%, **Figure 11, 12**).

**Figure 11.**
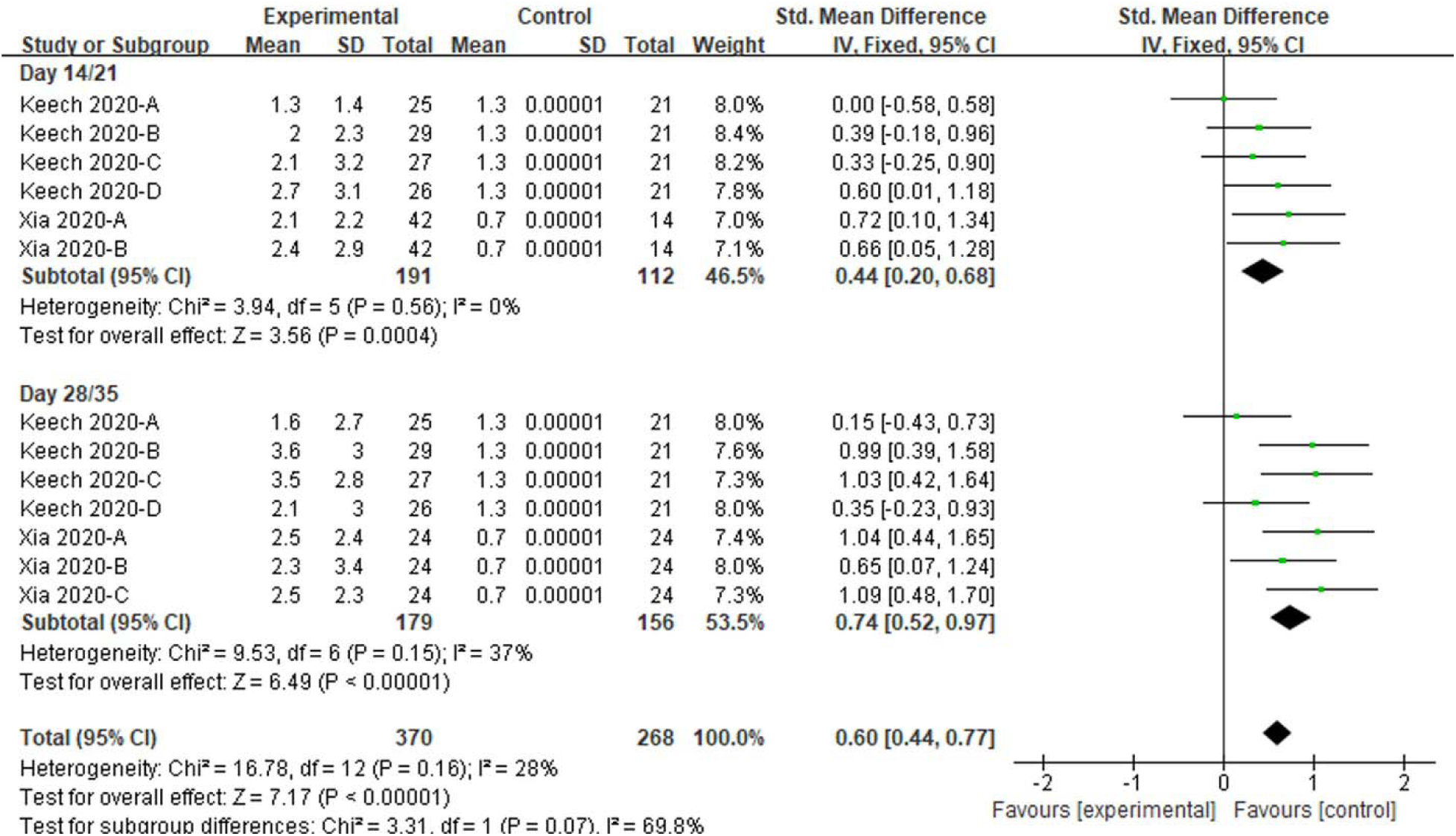
Meta-analysis of effect of vaccination on neutralizing antibody responses between before and post vaccine. Keech 2020-A: 25 µg rSARS-CoV-2/0 µg Matrix-M1 on first vaccination; 25 µg rSARS-CoV-2/0 µg Matrix-M1 on second vaccination; Keech 2020-B: 5 µg rSARS-CoV-2/50 µg Matrix-M1 on first vaccination; 5 µg rSARS-CoV-2/50 µg Matrix-M1 on second vaccination; Keech 2020-C: 25 µg rSARS-CoV-2/50 µg Matrix-M1 on first vaccination; 25 µg rSARS-CoV-2/50 µg Matrix-M1 on second vaccination; Keech 2020-D: 25 µg rSARS-CoV-2/50 µg Matrix-M1 on first vaccination; 0 µg rSARS-CoV-2/0 µg Matrix-M1 on second vaccination. Xia 2020-A: low dose vaccine at day 14 or 28 follow-up, Xia 2020-B: high dose vaccine at day 14 follow-up or medium dose vaccine at day 28 follow-up, Xia 2020-C: high dose vaccine at day 28 follow-up.

**Figure 12.**
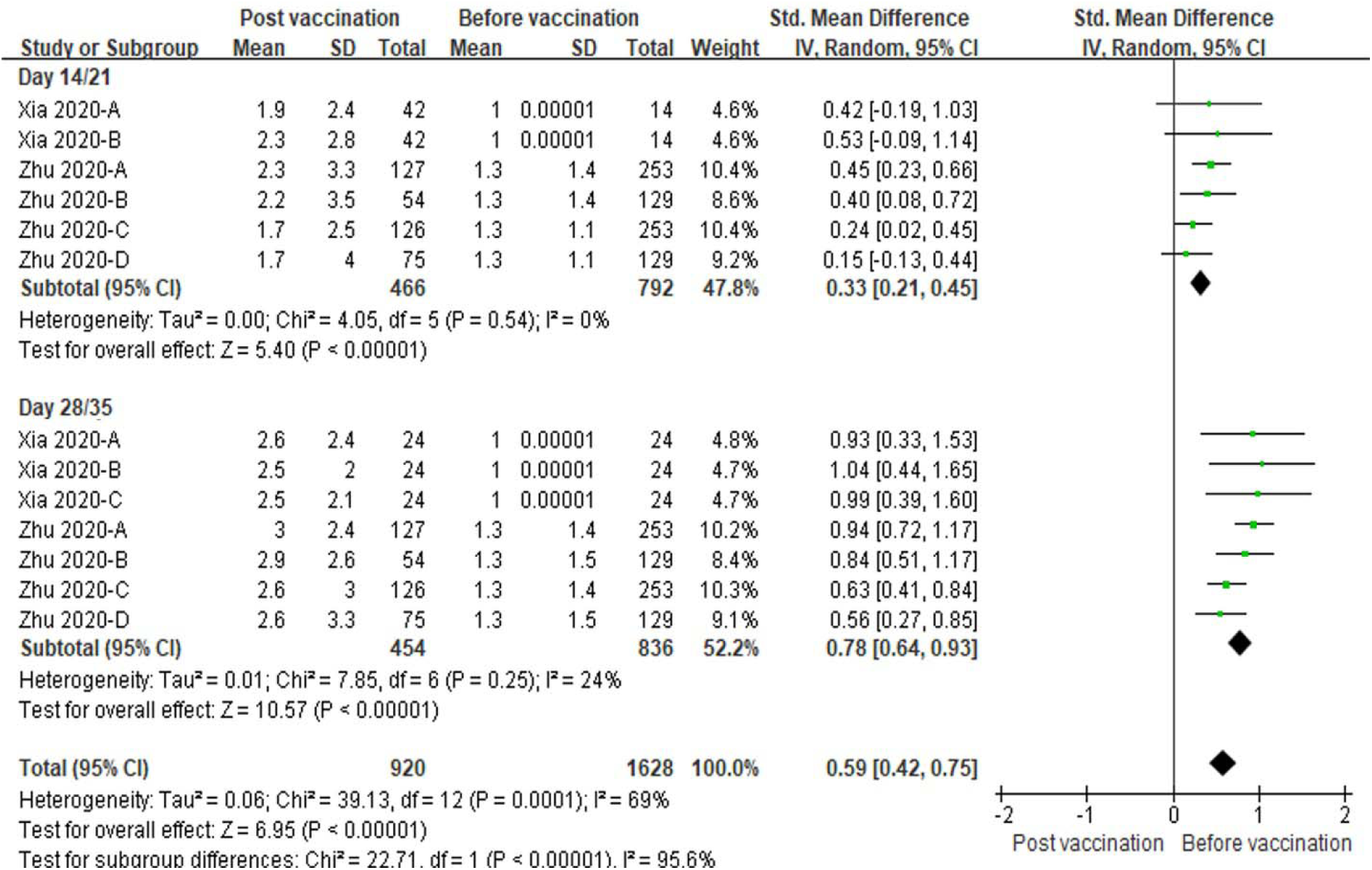
Meta-analysis of effect of vaccination on specific and IgG antibody responses between before and post vaccine. Xia 2020-A: low dose vaccine at day 14 or 28 follow-up, Xia 2020-B: high dose at day 14 follow-up or medium dose vaccine at day 28 follow-up, Xia 2020-C: high dose vaccine at day 28 follow-up. Zhu 2020-A: low dose vaccine and pre-existing Ad5 ≤ 200 geometric mean antibody titre. Zhu 2020-B: low dose vaccine and pre-existing Ad5 > 200 geometric mean antibody titre. Zhu 2020-C: low dose vaccine and pre-existing Ad5 ≤ 200 geometric mean antibody titre. Zhu 2020-D: low dose vaccine and pre-existing Ad5 > 200 geometric mean antibody titre.

### Effect of vaccination on laboratory parameters

In the laboratory parameters, the hemoglobin, alanine aminotransferase and total bilirubin in subjects with vaccine groups were similar as placebo groups (All *P* > 0.05, **Figure 13**). No amount of heterogeneity were identified related to three laboratory parameters in the analyzing studies (All I^2^ = 0%, **Figure 13**).

**Figure 13.**
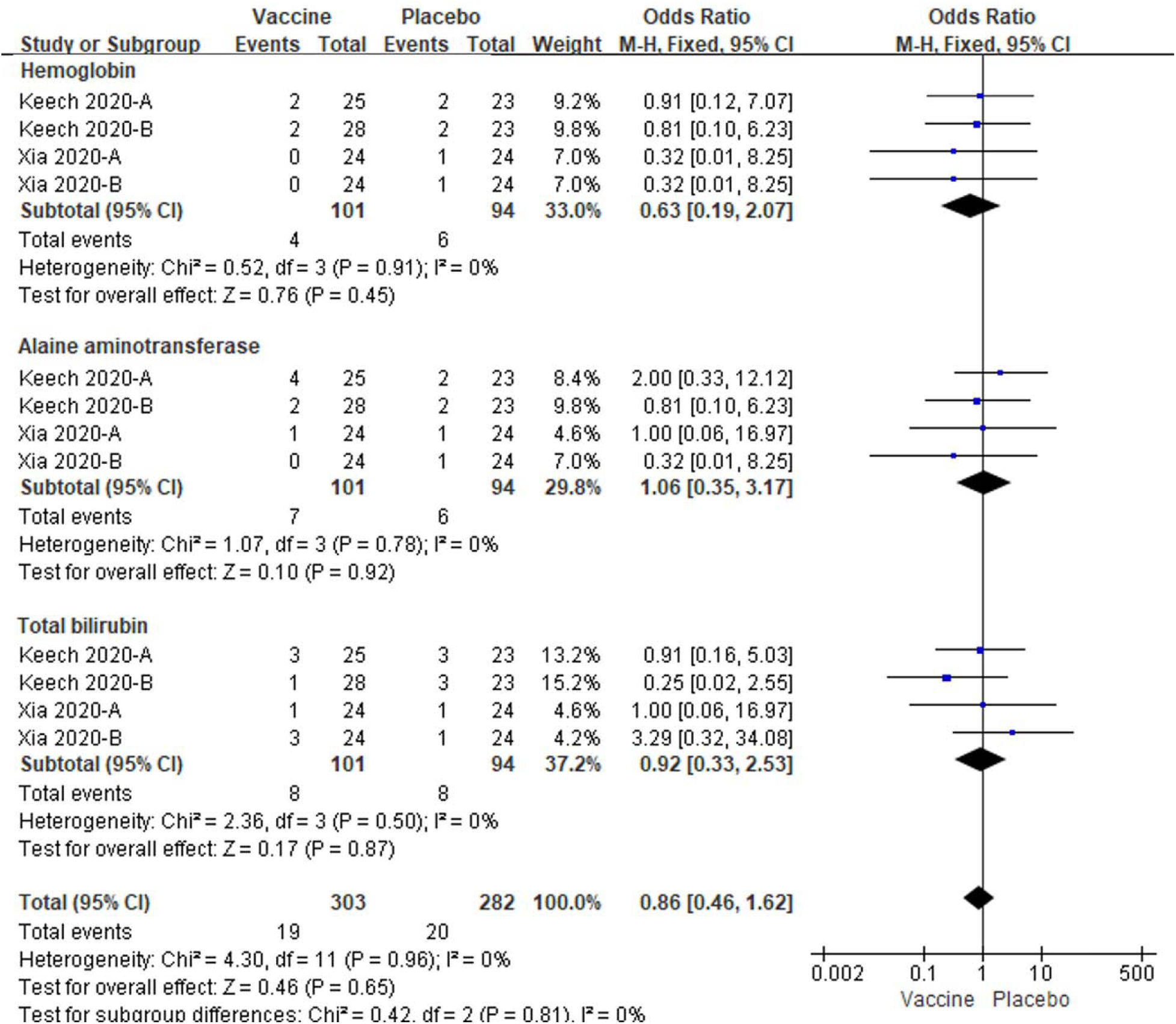
Meta-analysis of effect of vaccination on laboratory parameters between vaccine and placebo groups. Keech 2020-A: 25 µg rSARS-CoV-2/0 µg Matrix-M1 on first vaccination; 25 µg rSARS-CoV-2/0 µg Matrix-M1 on second vaccination; Keech 2020-B: 25 µg rSARS-CoV-2/50 µg Matrix-M1 on first vaccination; 0 µg rSARS-CoV-2/0 µg Matrix-M1 on second vaccination. Xia 2020-A: low dose vaccine, Xia 2020-B: high dose vaccine.

## Discussion

Immunotherapy is considered as an effective strategy for the prophylaxis and treatment of various infectious diseases, which requires the artificial triggering of the immune system to eliminate pathogens [12].

A vaccine that elicits the production of S protein neutralizing antibodies in the vaccinated subjects is the eventual aim for the all efforts that are devoted in the development of COVID-19 vaccine. To achieve this aim, multiple strategies have been applied. The most straightforward approach in vaccine production is to assemble SARS-CoV-2 S protein or its subunits with adjuvants directly. By this strategy, COVID-19 vaccines to elicit neutralizing antibody and immune cell (e.g. TH1 T cell) responses have been produced by multiple programs worldwide [13-15]. Another option is to utilize nucleoside-modified RNA (modRNA) including BNT162b1 [4, 16] and BNT162b2 [17] that encodes the SARS-CoV-2 receptor-binding domain (RBD) and the full-length S protein with 2 proline mutations for maintaining the prefusion conformation, respectively, as vaccine candidates. Besides, other types of vaccines including DNA-based vaccine candidate targeting SARS-CoV-2 S protein [18] and live attenuated vaccines with NS1-deleted RBD domain [19] has also been generated and examined.

Here, we have discussed data from 5 randomized, double-blind, placebo-controlled studies including 1246 cases received COVID-19 vaccine candidates and 358 subjects received placebo controls. So far, although it is still a small number for meta-analysis, it is the first meta-analysis to examine the safety, tolerability, and immunogenicity of current COVID-19 vaccine candidates. In the present study, the following conclusions were drawn.

1. Generally, to all analyzed vaccine candidates in our study, the majority of adverse events reported were mild or moderate in severity and no serious adverse reactions were reported. Additionally, all adverse effects were transient and resolved within a few days. However, it is important to emphasize that, no matter low or high dose of vaccination, vaccine recipients showed a significant increase in total adverse event versus controls. Furthermore, this elevation in total adverse event is mainly contributed by the significant increase of local adverse reactions. The total systemic adverse reactions displayed no significant difference among individuals vaccinated with or without COVID-19 vaccine, although some subdivision parameters may exhibit significant alterations. The total events of systemic reactions in low and high dose vaccination in Figure 6 and 8 were not consistent to the results in Figure 3 and 4. One main reason was that the total events in Figure 6 and 8 was only pooled some items at one follow-up timepoint and one dose injection not all the systemic reactions observed in studies, so they were not same with the results of polled all items about systemic reactions in Figure 3 and 4. Therefore, based on current observations, the COVID-19 vaccine candidates are overall safe and tolerated for clinical application.
2. All vaccine candidates exhibit excellent immunogenicity. 2-5 weeks post vaccination, both neutralizing and IgG antibody responses against SARS-CoV-2 S protein increased significantly in convalescent plasma samples. The specific immune responses against SARS-CoV-2 were constantly observed in individuals who received COVID-19 vaccine candidates, compared with no matter placebo-controls or the baseline before vaccination.
3. Multiple vaccinations, but not injection dose, are a key factor that determines the immunogenicity of current COVID-19 vaccine candidates. Multiple vaccinations of COVID-19 vaccine candidates always show greater immunogenicity than single vaccinations. In two included studies that give 2-3 injections, multiple vaccinations induced an increase around 10-100 times greater than single ones. When compared with convalescent serum, repeated vaccinations may result in GMT levels multiple folds greater than those in symptomatic outpatients with COVID-19 and approach comparable levels to those in hospitalized patients with COVID-19. In contrast, our analyses suggest that both low and high dose of vaccination can induce promising neutralizing and IgG antibody responses against SARS-CoV-2.

Therefore, meta-analysis results indicate great achievements in the development of safe, tolerated, immunogenic vaccines against SARS-CoV-2. However, this review has several limitations.

1. Some of the included studies exposed some methodological flaws, thereby introducing high risk of biases into these trials, that is, some trials failed to include the subjects at greatest risk for serious COVID-19, and outcome assessors after long-term period of follow-up. Moreover, some trials included insufficient number of samples without the wide ethnic and age varieties, which meant there was a potential risk of overestimating positive outcomes. Thus, further studies are required to increase the size of participant numbers, extend the follow-up time from 4-5 weeks to at least 3-4 months, and consider more population factors including adding older adults, people with coexisting conditions, and individuals with ethnically and geographically diversity.
2. We only include randomized, double-blind, placebo-controlled studies. Data from these articles can generate the most unambiguous results; however, there are multiple single-blind or placebo-free studies that are excluded unfortunately. For example, Sahin et al. reported that BNT162b1 elicited robust CD4^+^ and CD8^+^ T cell responses and strong antibody responses in a placebo-controlled, observer-blinded phase I/II vaccine trial [16]. Similarly, Folegatti et al. also demonstrated ChAdOx1 nCoV-19 with SARS-CoV-2 S protein as a safe, tolerated, and immunogenic vaccine in a single-blind, randomized controlled trial [14]. The studies on non-human mammalian species have also been excluded although [13, 15, 20]. These studies obtained promising results in animals in the perspective of immunogenicity and lung protection through using replication-incompetent recombinant serotype 5 adenovirus carrying a codon-optimized gene encoding S protein [20], insect cell-produced pre-fusion trimer-stabilized SARS-CoV-2 S protein [15], and CHO-expressed SARS-CoV-2 S1-Fc fusion protein [13]. We are aware of 5 includes was restricted to published studies and may therefore be affected by publication bias. Thus, It is urgently needed to carry out more randomized, double-blind, placebo-controlled studies, include a large number of studies in a broad geographical scope, in order to obtain a more comprehensive view of the safety, tolerability, and immunogenicity of current COVID-19 vaccine candidates as a result.
3. Another one is the lack of some key data since detailed patient information was not given in all studies, especially regarding to SARS-CoV-2 anti-Spike (S) protein IgG and neutralizing antibody responses, and biochemical indicators of immune responses that are obtained by blood routine, blood biochemical, urine routine, and other test methods. To examine the complete physiological and biochemical indexes of all participants should be an important part in following trials.
4. Although there was no time restriction in literature search, we only collected English language papers. Due to the possibility that the language restriction may narrow the breadth of our search, more languages (e.g. Chinese, Spanish, Japanese, French, and Russian) and database (e.g. MEDLINE, Ovid MEDLINE In-Process, and Scopus) should be applied in literature search in the following studies.

In summary, our study is the first meta-analysis summarizing the current progress in the SARS-CoV-2 vaccine development. The results from our analysis implicate excellent safety, tolerability, and immunogenicity of all included COVID-19 vaccine candidates, providing great confidence for the following development and clinical application of COVID-19 vaccines. Therefore, our study suggests that, although there are limitations in current studies such as the short follow-up time and small sizes of subjects, with the conduction of large-scale clinical efficacy trials, the successful development of safe, effective, durable, and deployable to large populations COVID-19 vaccines to end the global SARS-CoV-2 pandemic will be a near possibility.

## Data Availability

All datasets generated for this study are included in the manuscript.

## Declarations

### Ethical Approval and Consent to participate

Not applicable.

### Consent for publication

The authors approved the final manuscript.

### Availability of supporting data

Not applicable.

### Authors’ contributions

J.C.Z., X.X. conceived the manuscript. P.Y., P.A., Y.H., X.X. collected references. P.Y., P.A., Y.H., Z.A. analyzed the data. P.Y., P.A., Y.H., X.X. wrote the manuscript. All authors reviewed and approved the final article.

## Acknowledgements

We thank Jie Zhu, Yanyan Zhang, Drs. Ling Ye and Xinrui Qi for proofreading the manuscript.

